# Remote-management of COPD: Evaluating Implementation of Digital Innovation to Enable Routine Care (RECEIVER) – Protocol for a feasibility and service adoption observational cohort study

**DOI:** 10.1101/2021.02.04.21251162

**Authors:** A Taylor, G McDowell, S Lua, S Burns, P McGinness, D Lowe, C Carlin

## Abstract

**Introduction:** Reduction in exacerbations and hospitalisations are the outcomes rated as most important by COPD patients. Patient self-management and evidence-based healthcare interventions that reduce exacerbations and hospital admissions do exist but there are considerable barriers to their uptake and delivery. Patient and clinician engagement is often not consistent. Data which could potentially predict exacerbations and treatment success are not acquired systematically and is not reliably visible or actionable at key time points of patient-clinician interaction. Most COPD management is currently based on a reactive approach, and delays in recognising treatable opportunities underpin COPD care-quality gaps. Innovations which can empower patient self-management, facilitate integrated clinical care and support delivery of evidence-based treatment interventions are urgently required.

We have developed a digital service model for COPD patients, designed to integrate current routine clinical care within a digitally enabled remote-monitoring service infrastructure. This digital platform will capture relevant routinely acquired patient-reported outcomes, continuous physiology data and clinical event/episode data in a patient and clinician co-designed interface.

**Methods and analysis:** The remote management of COPD: evaluating implementation of digital innovations to enable routine care (RECEIVER) trial is a prospective observational cohort hybrid implementation and effectiveness study that will test the adoption of this digital service model for ‘high-risk’ COPD patients and evaluate the performance of this approach versus current standards of care. Patients with a recent severe exacerbation and/or COPD-OSA overlap or chronic hypercapnic respiratory failure requiring home NIV or CPAP, with access to smartphone, tablet or computer will be recruited into the study and enrolled into the digital service.

In our endpoints will determine patient engagement, clinical service impact and clinical outcomes that will be compared with historical and contemporary COPD control patient data, acquired from NHS GG&C SafeHaven. The digital innovations will be iterated throughout the study to optimize them, based on the user experience accrued. The digital infrastructure for this support of routine clinical care will also provide a foundation to explore the feasibility of approaches to predict outcomes and exacerbations in COPD patients, for testing in future prospective clinical and regulatory trials through machine-learning analysis.

**Ethics and dissemination:** Ethical approval for this clinical trial has been obtained from the West of Scotland Research Ethics Service (WoSRES). The trial will commence in September 2019 for a duration of 2 years. Key results will be presented at local, national and international meetings, including those with patient representation. All data obtained will be submitted for publication to peer reviewed journals.

**Trial registration number:** NCT04240353

## Introduction

Chronic obstructive pulmonary disease (COPD) is a common, preventable and treatable disease, characterised by persistent respiratory symptoms and airflow limitation that is due to airway and/or alveolar abnormalities. The main risk factor for COPD is tobacco smoke but other environmental exposure may contribute.

The most common respiratory symptoms are breathlessness, cough and/or sputum production. These symptoms may be under-reported by patients. COPD may be punctuated by periods of acute worsening of respiratory symptoms, often referred to as exacerbations, which can result in emergency department attendance or hospital admission. For many patients, COPD is associated with significant comorbidity, which increases its morbidity and mortality.

COPD is a major healthcare challenge, with worldwide rising prevalence. The Global Burden of Disease Study reported a prevalence of 251 million cases of COPD globally in 2016^1^. It is projected to be the 4th leading cause of death worldwide by 2020^2^. Reductions in exacerbations and hospitalisations are the outcomes rated as most important by COPD patients^3^. Effective delivery of evidence-based interventions for COPD - smoking cessation, influenza vaccination, pulmonary rehabilitation, personalised inhaled therapy, home oxygen therapy (where indicated) and home non-invasive ventilation (NIV, where indicated) – have been shown to reduce exacerbations and hospital admissions^4^. There are considerable barriers to uptake and delivery of evidence-based interventions^5^. This care-quality gap particularly affects outcomes from COPD exacerbations. COPD exacerbations are the main driver of healthcare costs (estimated annual NHS cost of managing COPD is £1.9bn^6^). Service redesign, based on innovations that can integrate care to deliver these evidence-based interventions to address this care-quality gap and achieves reduction in COPD exacerbations and admissions, is urgently required.

Self-management also plays a key role in the treatment of COPD^7^. Patients who can be successfully taught and supported with COPD self-management show a significant reduction in COPD admissions^8,9^. The establishment of multi-disciplinary community respiratory team in NHS Greater Glasgow & Clyde (NHS GG&C), supporting self-management in patients identified acutely as being high-risk of hospital admission, has been associated with reduction in hospital admission rates^10^.

Whilst interventions should be developed for patients with all severities of COPD, it is logical to target immediate efforts towards patients with ‘high-risk’ COPD, i.e. those who are at most risk of exacerbations and hospital admissions. Established data indicates that COPD patients who have had a severe exacerbation (one requiring emergency department attendance or hospital admission) in the previous 12 months and/or have persisting hypercapnic respiratory failure fall into this high-risk group^4,11^. Interventions proven in this group can then be rolled out (if cost-effective) to the lower-risk groups of COPD patients.

### Digital Service Model

Pilot data from NHS GG&C has highlighted the potential for digital innovations to predict COPD outcomes and support treatment uptake. Slevin et al. interviewed patients with COPD and showed their wiliness to take a more active role in self-management using digital health technology (DHT), along with their acknowledgement of how DHT could play a role in supporting healthcare professionals to practise preventative care provision^12^. Web and smartphone-based apps have shown the capability to facilitate disease self-management and support uptake of interventions^13,14^ (ref). Although COPD patient focused digital tools currently exist, there is a limited evidence-base for their use, with evaluations mainly performed in isolation and no integration with established clinical services or statutory electronic health records.

Home non-invasive ventilation (NIV) successfully improves admission-free survival, in patients who have persisting hypercapnic respiratory failure following a life-threatening COPD exacerbation, with a number needed to treat of 7 patients^15^. There has so far been relatively limited success at providing home NIV for COPD patients in the UK, within existing service models. However, in NHS GG&C we have been successful in delivering home NIV to eligible COPD patients, utilising digital technologies routinely available (adaptive “auto-NIV” modes, 2-way remote monitoring via ResMed AirView platform), with outcomes in our service adoption pilot study matching those from the HOT-HMV randomised clinical trial^16,17^. The challenge is to extend the evidence for this approach and obtain a service adoption playbook to enable this to be adapted and delivered at scale, by other clinical teams, within COPD integrated care.

Most COPD management is currently based on a reactive approach, and delays in recognising treatable opportunities underpin COPD care-quality gaps. For example, patients with a COPD exacerbation typically have symptom deterioration for 2 days before seeking assistance, and then a potential 2 to 5-day delay in accessing routine schedules clinical care. Several studies have indicated the ability of regularly recorded patient-reported outcomes and physiology measurements to predict outcomes, including exacerbations and treatment success/failure in COPD patients. Changes in COPD symptom diary scores and home NIV parameters including respiratory rate can predict COPD exacerbation development^18,19^. Changes in activity measured by wearable devices predict outcomes after COPD exacerbations^20^. Currently symptom diaries and other patient reported outcome questionnaires, activity, exercise and NIV data are obtained in routine practice in NHS GG&C. However, patient and clinician engagement are not consistent, data are not acquired systematically and are not often visible or actionable at key time points of patient-clinician interaction. These shortfalls, and the arising care-quality gaps, could potentially be addressed by digitising this routine clinical care, improving the patient clinician interface for data entry and collating the acquired data.

Patient-clinician communication for COPD management, including supporting self-management, is currently dependant on face-face scheduled consultations, answerphone/email asynchronous messages from patient > clinician, and unstructured advocacy triggered or initiated communication from clinician > patient. These present several inefficiencies and risks, which could be overcome by digitising their patient-clinician messaging system to support scheduling, remote management and support COPD self-management.

Machine-learning analysis and modelling based on available data shows significant promise in COPD predictive management. Data available in patient’s electronic health record (EHR) at triage assessment can robustly predict outcome (admission, length of stay) from a severe COPD exacerbation^21,22^. The addition of physiology measurements to EHR data improves machine-learning predictive model performance in other clinical conditions^23^. Further evaluation of these analytics and predictive modelling, in a comprehensive dataset including patient-reported outcomes, physiology data and clinical events is a logical step to determine their potential role in real-time or near real-time clinical use.

### Rationale

Innovations which can empower patient self-management, facilitate integrated clinical care and support delivery of evidence-based treatment interventions are urgently required. In the RECEIVER trial, we propose to test the implementation of a platform which digitises these as an additional – potentially assistive – component alongside routine clinical care. In our endpoints will determine patient engagement, clinical service impact and clinical outcomes, to evaluate the performance of this approach versus current standards of care. A digital infrastructure for this support of routine clinical care would also provide a foundation to explore the feasibility of approaches to predict outcomes and exacerbations in COPD patients, for testing in future prospective clinical and regulatory trials.

Our aims are to establish a digitised service model for ‘high-risk’ COPD patients which will:

– Integrate current routine clinical care within a digitally enabled remote-monitoring service infrastructure
– Enable delivery of remote management of COPD at scale within the NHS and other healthcare systems
– Capture relevant routinely acquired patient-reported outcomes, continuous physiology data and clinical event/episode data in a patient and clinician co-designed interface which enables engagement
– Facilitate evolution from a reactive to a proactive and preventative COPD service model

Key components of proposed digital service model for COPD are noted, and summarised in Figure 1. Appendix 2 provides a table outlining how the RECEIVER trial components compare with current routine clinical care.

**Figure 1.**
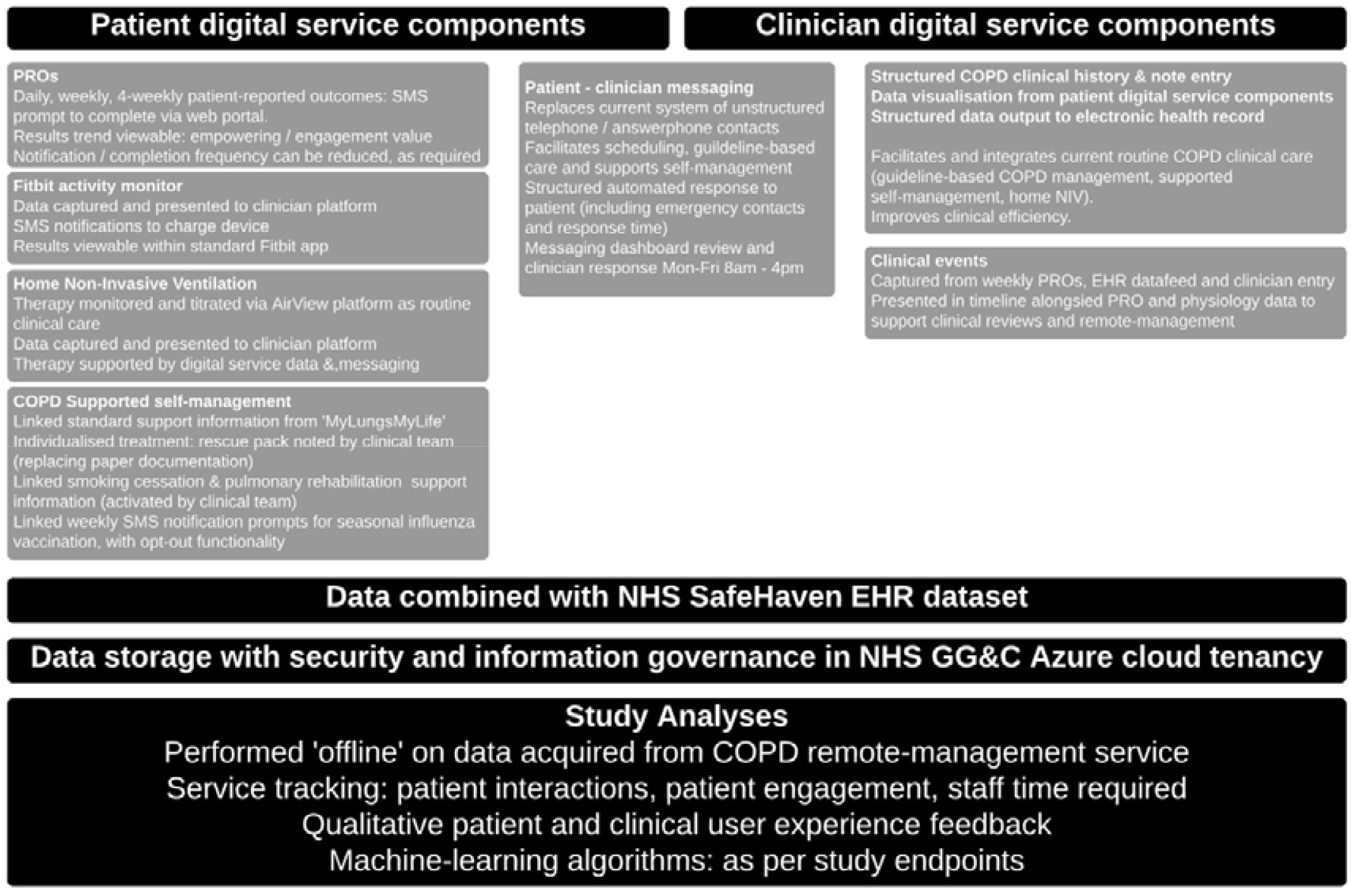
RECEIVER Digital Service Components.

The COPD digital service components being utilized in the RECEIVER trial are:

– **Patient facing web portal**: This has been co-designed with COPD patients and captures patient reported outcomes (PROs) and provides access to standard COPD self-management content. It also includes a messaging facility which can be patient, or clinician initiated. Examples of visuals available on supporting website, https://support.nhscopd.scot
– **Patient wearable device**: Fitbit Charge 3 device (CE marked)
– **Remotely monitored home NIV**: provision of this is our current standard of care for patients with hypercapnic respiratory failure, using AirView NIV remote management platform (ResMed). We have integrated our COPD digital service platform with the AirView platform, so that it acquires the NIV data unmodified for review. NIV therapy management will continue to be conducted through the AirView platform: the RECEIVER trial COPD platform is not used to modify or enhance patient NIV therapy.
– **User designed clinical dashboard:** presents integrated data with an aim of facilitating improvement in provision of guideline-based COPD care, and supporting COPD self-management.
– **Patient**-clinician asynchronous messaging: to support routine clinical care.
– **NHS GG&C Azure cloud-based digital architecture (appendix 3):** this provides and integrates the above services with existing NHS GG&C electronic healthcare systems.

## Study Methods and Analysis

### Study Design

This study is a prospective observational cohort hybrid implementation and effectiveness study^24^. It will be performed according to the UK Policy Framework for Health and Social Care Research (2020)^25^. The clinical intervention component is regarded as a phase IV adoption study. Data visualization to facilitate guideline-based care, supported self-management and home NIV are evidence-based COPD interventions.

In this study, we will evaluate the adoption of digitally integrated remote-management service innovations to support routine clinical care. Implementation of these will be evaluated and the digital innovations will be iterated to optimize them, based on user experience accrued during the study. We will acquire a consented dataset for trial endpoint analysis including exploratory machine-learned predictive modelling. The machine-learning analysis will also allow us to priorities data inputs for follow up study.

Outcomes and other variables in the prospective cohort will be compared with historical and contemporary COPD control patient data, acquired from NHS GG&C SafeHaven.

We will conduct a sub-study of additional baseline and follow up physiological measurements (oscillometry, parasternal EMG, home pollution monitoring) in patients in whom it is feasible to obtain these during their hospital admission and/or hospital attendance alongside their routine clinical care. We will also conduct a sub-study with consented digital service clinical users, with platform tracking analytics to measure clinician time spent on platform components, and qualitative user experience data acquisition (presented spontaneously and in semi-structured group or 1:1 interviews).

An overview of the study design is provided in Figure 2.

**Figure 2.**
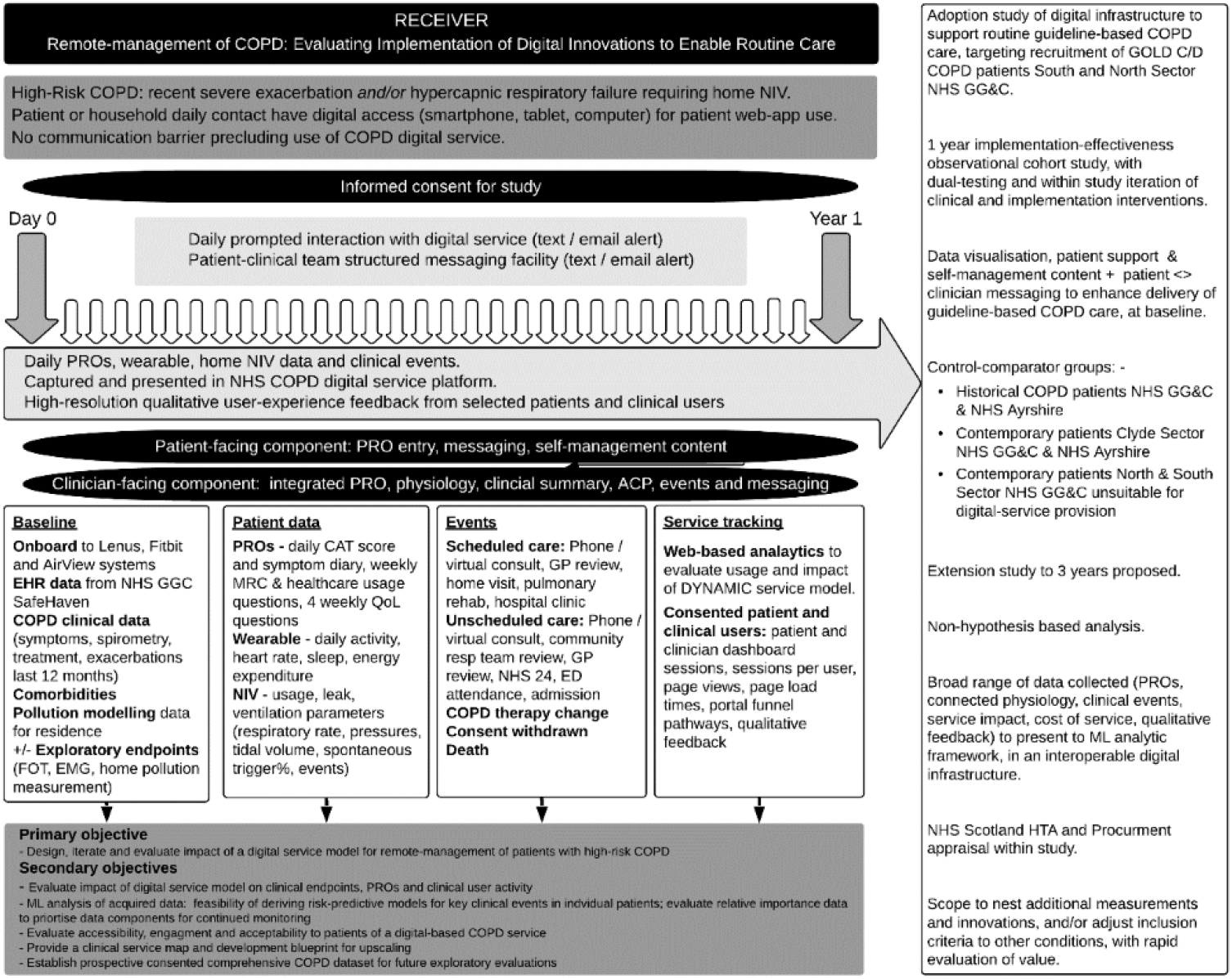
RECEIVER study design.

### Study Population

Patients will high-risk COPD attending secondary care in North and South Sector of NHS GG&C will be screened for inclusion in this study.

#### Inclusion Criteria

- Confirmed diagnosis of chronic obstructive pulmonary disease, established pre-screening or at screening, defined as per GOLD 2019 guidelines^4^
- Home non-invasive ventilation cohort: hypercapnic respiratory failure and/or sleep disordered breathing, meeting established criteria for provision of home NIV
- Exacerbation cohort: presentation within last 12 months with severe or life-threatening exacerbation of COPD, defined as per GOLD 2019 guidelines^4^
- Patient or close-contact who has daily access to smartphone, tablet or home computer with broadband access to web-browser
- Informed consent
- ≥18 years of age

#### Exclusion criteria

- Inability to comprehend informed consent
- Communication barrier precluding use of COPD digital service

### Sample Size

Previous NHS GG&C studies^8,16^, combined with internal audit data (indicating that 80% of patients have direct or daily smartphone, tablet, or web access) indicate that recruitment of 400 patients across a 12-month period is feasible, and likely to provide adequate data to conduct meaningful primary and secondary endpoint analyses.

An improvement in admission-free survival, with reduction in admissions by 1 per patient per year in projected, if support for provision of evidence-based COPD care is achieved by the service model. As feasibility of an end-end digitised service model for COPD has not yet been tested, and the service components will be adapted during the study, sample size power calculations and randomisation are not yet appropriate.

### Screening and eligibility assessment

Patients will be identified from emergency department (ED) attendance, hospital admission or COPD multi-disciplinary team referral. It is anticipated that the majority of patients will be identified and recruited during their index acute admission.

Review of focused medical history and lung function test results will be undertaken as part of screening evaluation to ensure all inclusion and no exclusion criteria are met.

### Informed consent

We will undertake informed consent with timing which is individualized to be least burdensome and most efficient for patients. Written informed consent will be obtained by participant’s dated signature and dated signature of the person who presented and obtained the informed consent. The person who obtained the consent will be suitably qualified and experienced and have been given authority to do so by the principle investigator.

In addition to consent for the clinical trial, GDPR consent for data sharing, data storage and tracking of web-app activity is required, and this is sought within the COPD patient-app when patients complete initial sign-up to the service.

### Withdrawal of subjects

Participants have the right to withdraw from the trial at any point, for any reason, without prejudice to future care. This is clearly stated at time of consent. The investigator can also withdraw patients from the study intervention in the event of protocol violations or any other relevant reasons. Patient withdrawals will be included within the study analyses and reporting.

Patients may opt to limit the number of monitoring interventions – e.g. daily questionnaires that they undergo, if these are burdensome.

Patients may also request ‘conventional’ scheduled face-face clinical review rather than remote monitored review at any time point. This will be accommodated as outpatient or inpatient attendance, within routine clinical timescales as judged appropriate by the clinical team, in-line with current service provision.

### Patient identification

Study numbers will be assigned sequentially as each subject enters the study to ensure that study patient data is de-identified. Corresponding information will be recorded on the Case Report Form (CRF) by the investigator.

### Source data

Source documents will be the hospital medical records, clinical charts, laboratory and pharmacy records, radiographs and correspondence. COPD digital service web-portal and clinician dashboard will be the source data for clinical summary data, patient-reported outcomes, wearable and NIV physiology, patient-clinician messaging, clinical documentation and exacerbation / admission recording in participants. The patient and clinician dashboard will also output this source data as structured report to NHS GG&C clinical portal (patient’s electronic health record).

### Study data

Electronic data and COPD service digital architecture will be held within NHS GG&C e-health systems, with industry standard security and identity assurance processes. The core components of digital service will be on NHS GG&C Azure cloud tenancy, further ensuring security and identity assurance, and avoid need for data de-identification prior to machine-learning analysis.

Data access will be password protected and accessible only by study investigators, with data management as per NHS GG&C and NHS Scotland data protection policies. Platform analytics will track user interactions and patient data changes, to provide audit trail for data integrity.

### Subsequent assessments

Participants will be recruited into the digitalised service model over a 12-month period. Observation of secondary endpoints will be continued over the 12-month period following recruitment. Patients recruited will subsequently continue within the digital service model beyond the trial period, either until ongoing procurement of the service infrastructure is established, or transition to alternative equivalent service model is completed.

### Definition of end of trial

The end of trial will be when the last participant has completed follow up, 12 months after recruitment.

### Patient and public involvement

Patients and members of the public were involved throughout the planning and conduct of this research. The patient-facing web application, which forms the basis of the study, was co-designed with COPD patients and carers. Semi-structured interviews were conducted between patients and the development team to understand patient experience of their condition, with feedback and adaptation of the application at each development stage. Subsequent development of research questions and study design was informed by insights gained through these interactions.

## Data Acquisition

### Electronic Health Record Data

Demographic, coded diagnosis list, Charleson comorbidity index, Scottish Index of Multiple Deprivation quintile, medication history, laboratory results, lung function results, emergency department attendance, hospital admission, pulmonary rehabilitation attendance data from 1^st^ January 2010 onwards will be obtained from NHS GG&C SafeHaven, and presented for evaluation in study machine learning analysis platform.

### Baseline Clinical Data

Patient age, sex, height, weight, smoking status, vaccination status, comorbidities, COPD exacerbation history, COPD medications, inhaler technique, lung function results, key laboratory results including maximal eosinophil count and modelled home pollution exposure (QCumber platform; NO2, O3, PM2.5, Black carbon) will be documented.

Patients who have not had contemporary spirometry (standard lung function test) will have this repeated at time of recruitment – this would be routine clinical care.

A subset of patients (where time and mode of presentation allows) will be offered recruitment to the exploratory physiology sub-study and have additional physiology measurements take. These will comprise of oscillometry, parasternal EMG and home air pollution pack monitoring. Measurements from these may predict / associate with stability of COPD and clinical endpoints, and follow up measurements may be taken at 3, 6, 9 and 12 months if feasible during study, alongside routine clinical care. The principle purpose of conducting these in this study is to report on feasibility within routine COPD assessment.

### Follow-up Clinical Data

Smoking cessation, vaccination status, pulmonary rehabilitation, COPD comorbidity, COPD exacerbation history, COPD medications and other treatments, inhaler technique and narrative history will be updated at clinical encounters, as relevant.

Data feed from NHS GG&C Trakcare platform to RECEIVER study COPD platform will present information on hospital attendance and admissions.

Clinical episodes are also captured in the weekly patient-reported outcome questionnaires, and manually inputted by clinical team, when these are noted.

Accuracy of coding of clinical summary data and clinical episodes will be reviewed in a proportion of recruited patients, by a respiratory physician working independent of this study.

### Patient Reported Outcomes

Patients (or their family or carer) will complete patient reported outcome (PRO) questionnaires in the patient web portal. Daily text and/or email notifications are provided as a prompt to complete these. There is support information in the associated website to assist patients with any difficulties with the patient portal.

Patients will complete symptom diary and CAT questionnaire daily. MRC and healthcare episode questionnaires will additionally be completed weekly. EQ5-D quality of life questionnaire is additionally be completed every 4 weeks. These questionnaires have been integrated to improve question flow and simplify patient experience. Appendix 1 contains questionnaire flow content.

Pilot user experience research shows that the daily PRO questions can be completed in ∼70 seconds, with weekly questions taking an additional 90 seconds, and quality of life questions taking additional 5 minutes.

PRO data is presented unmodified, in a ‘user-friendly’ format (co-designed with clinical user), in the clinician dashboard. This unmodified data will inform and enhance clinical encounters and patient-clinician communication.

### Wearable Physiology Measurement

Patients will be provided with a study Fitbit Charge 3 wristband wearable device, to monitor activity, sleep, heart rate and energy expenditure variables. COPD patient portal will provide device support instructions.

Wearable physiology results will be available to patients in the Fitbit app.

Wearable physiology results are presented unmodified in study clinical dashboard, to inform and enhance clinical encounters and patient-clinician communication.

### Remote-Monitored Home Non-Invasive Ventilation

Patients with severe COPD who have standard indications (persistent hypercapnia with PCO2 >7kPa and/or recurrent episodes of acute hypercapnic respiratory failure and/or COPD-obstructive sleep apnoea overlap and/or COPD with significant nocturnal hypoventilation) are commenced on home non-invasive ventilation (NIV) in NHS GG&C using Resmed Lumis 150 ST-A device as routine clinical care. Patients with significant obstructive sleep apnoea syndrome may be alternatively commenced or transition to auto-titrating CPAP using AirSense-10 device. NIV initiation and optimization will be conducted as per standard NHS GG&C clinical protocol. NIV data capture and device adjustment will be via ResMed AirView platform, again as routine clinical care.

AirView data will be acquired via Open-API to the COPD platform, and presented in study COPD clinical dashboard, to improve data visualization for NIV clinical management alongside PROs and wearable data, and to inform and enhance other clinical encounters and patient-clinician communication.

### Patient-portal support materials, care plans, and clinical decision support

Patient platform will contain linked content to NHS GG&C smoking cessation, vaccination, inhaler therapy, pulmonary rehabilitation, breathing control and ‘my lungs, my life’ COPD support literature, all to aid self-management.

Patient-resource will contain a structured exacerbation self-management care plan. This is a digitalised version of the paper structured self-management plan without any change in format or content. Patients will be empowered to activate their own self-management plan or supported/directed to activate this with patient-clinician communication, as standard routine clinical care: the data access, visualisation and patient-clinician messaging facilities in this study will however enhance this.

### Machine-learning analysis and predictive modelling

These analyses will be conducted post-hoc, using the clinical data obtained from the RECEIVER trial COPD platforms, alongside EHR data in historical and contemporary control patients from NHS GG&C SafeHaven. The anticipation is that following analysis of the study data, we will be able to judge the feasibility and accuracy of utilising predictive model outputs at a service or individual patient level. This may lead to the subsequent development and evaluation of AI-based clinical decision support tools. Relevant clinical investigation/medical device trial(s) of these would be proposed and designed. The data from the RECIEVER trial would not be used directly to support any clinical validation of any subsequently developed medical device.

### Service model iterations

The components of the RECEIVER service model have been subject to pilot evaluation and pre-trial patient and clinical user experience research. Service refinement based on the clinical user sub-study experience research will continue during pre-trial preparation and approval period. Iterations of the digital service content (e.g. rationalisation of patient outcome, modification or addition of self-management support materials) based on adoption experience will be considered by the project steering group at 3 monthly intervals during the study. Where this iteration would result in a change in the clinical protocol, this will be submitted as an amendment for consideration and regulatory approval, before any change is implemented.

## Study Outcome Measures

Primary and secondary endpoints for RECEIVER study are shown in Table 1.

**Table 1:**
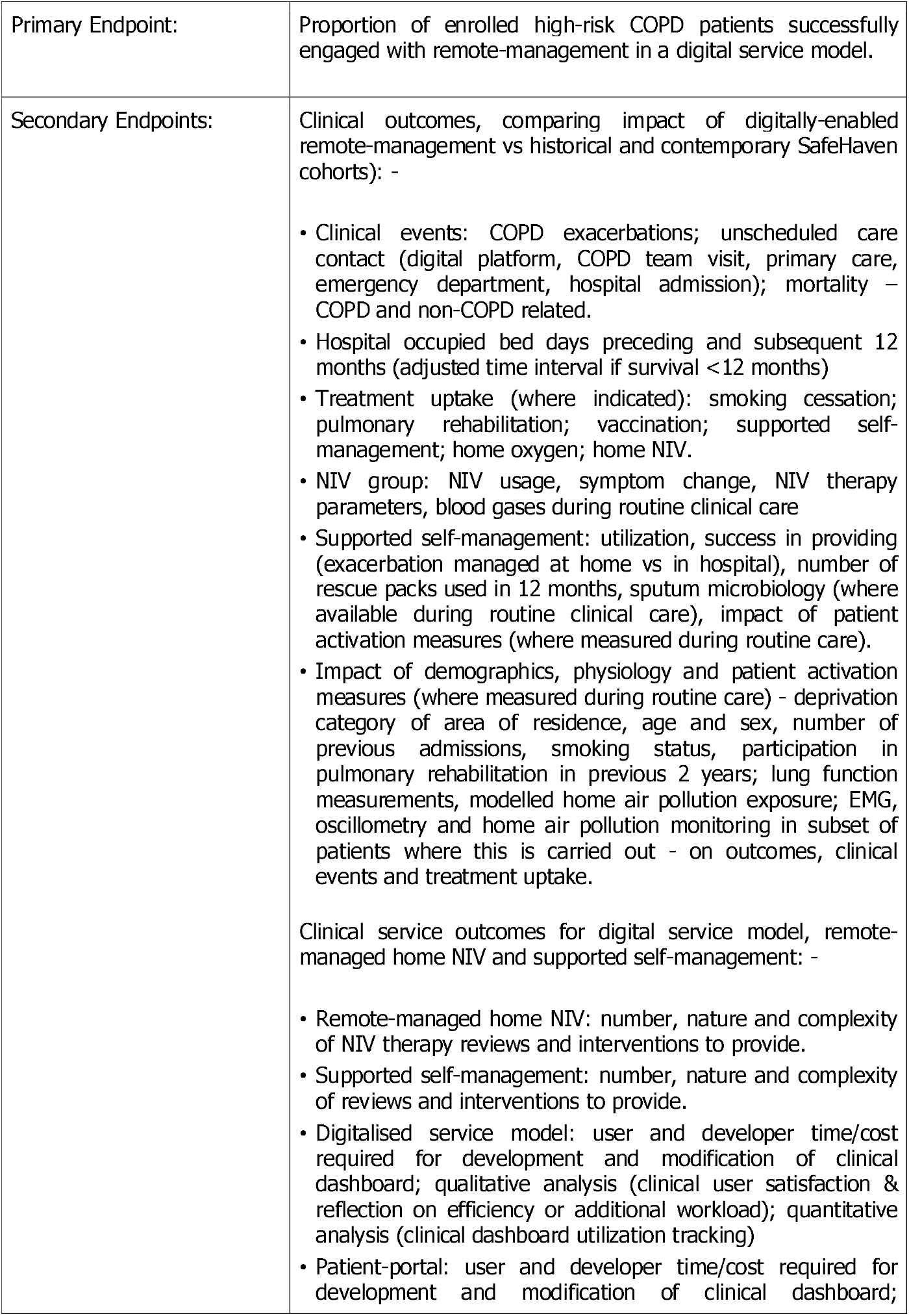
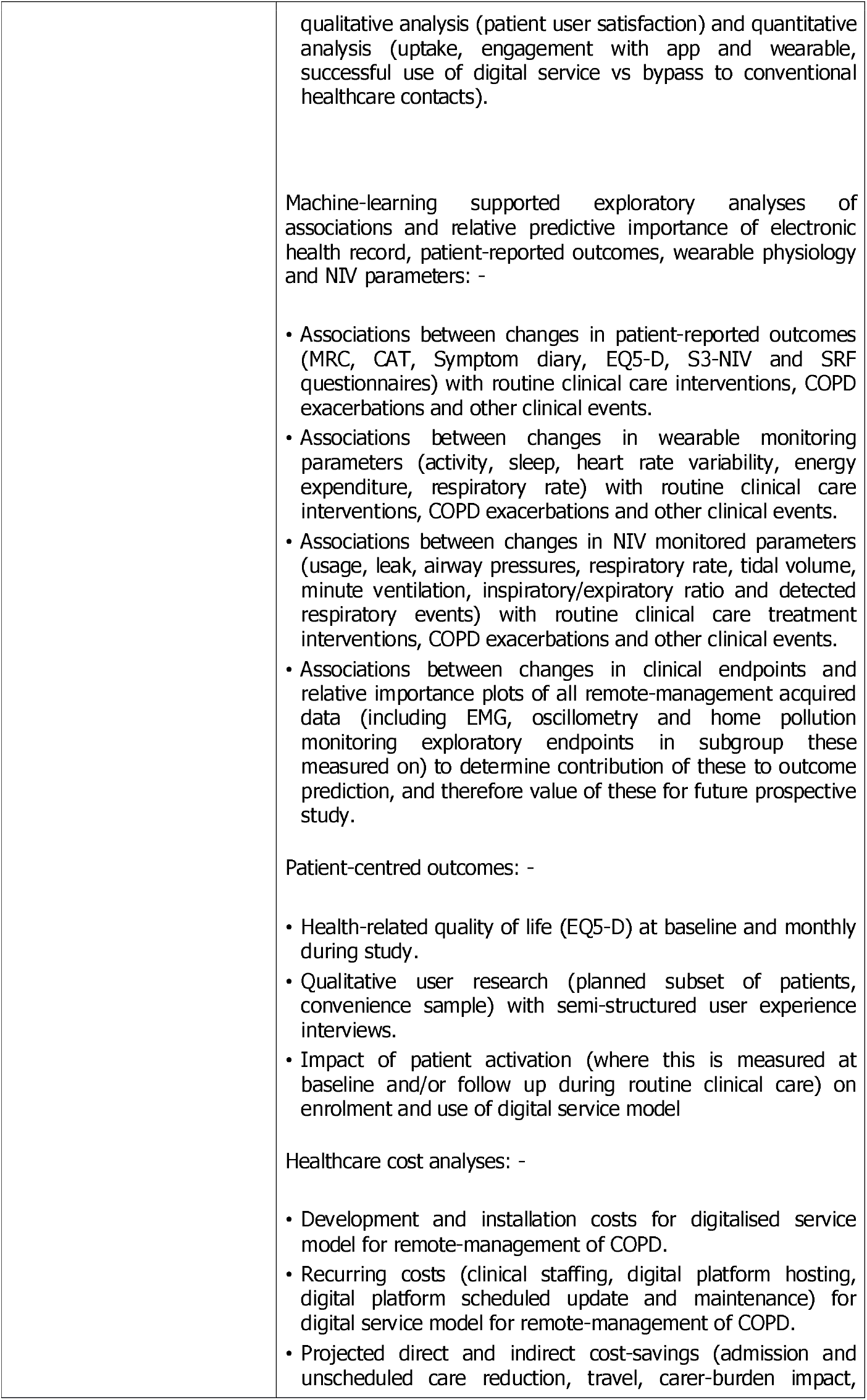
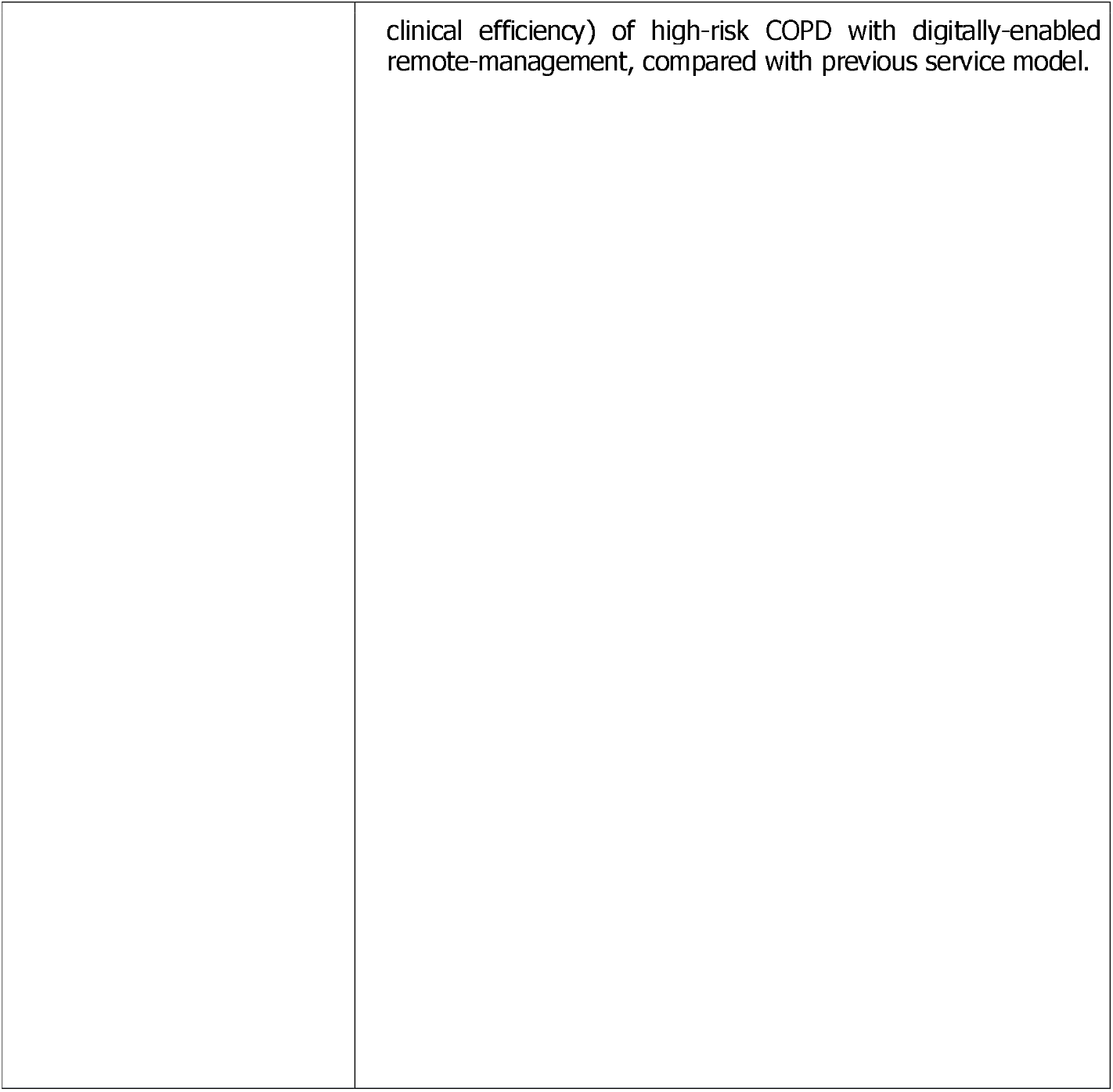
RECEIVER primary and secondary endpoints.

|  |  |
| --- | --- |
| Primary Endpoint: | Proportion of enrolled high-risk COPD patients successfully engaged with remote-management in a digital service model. |
| Secondary Endpoints: | Clinical outcomes, comparing impact of digitally-enabled remote-management vs historical and contemporary SafeHaven cohorts): -<br>• Clinical events: COPD exacerbations; unscheduled care contact (digital platform, COPD team visit, primary care, emergency department, hospital admission); mortality – COPD and non-COPD related.<br>• Hospital occupied bed days preceding and subsequent 12 months (adjusted time interval if survival <12 months)<br>• Treatment uptake (where indicated): smoking cessation; pulmonary rehabilitation; vaccination; supported self-management; home oxygen; home NIV.<br>• NIV group: NIV usage, symptom change, NIV therapy parameters, blood gases during routine clinical care<br>• Supported self-management: utilization, success in providing (exacerbation managed at home vs in hospital), number of rescue packs used in 12 months, sputum microbiology (where available during routine clinical care), impact of patient activation measures (where measured during routine care).<br>• Impact of demographics, physiology and patient activation measures (where measured during routine care) - deprivation category of area of residence, age and sex, number of previous admissions, smoking status, participation in pulmonary rehabilitation in previous 2 years; lung function measurements, modelled home air pollution exposure; EMG, oscillometry and home air pollution monitoring in subset of patients where this is carried out - on outcomes, clinical events and treatment uptake.<br>Clinical service outcomes for digital service model, remote-managed home NIV and supported self-management: -<br>• Remote-managed home NIV: number, nature and complexity of NIV therapy reviews and interventions to provide.<br>• Supported self-management: number, nature and complexity of reviews and interventions to provide.<br>• Digitalised service model: user and developer time/cost required for development and modification of clinical dashboard; qualitative analysis (clinical user satisfaction & reflection on efficiency or additional workload); quantitative analysis (clinical dashboard utilization tracking)<br>• Patient-portal: user and developer time/cost required for development and modification of clinical dashboard; |
|  | qualitative analysis (patient user satisfaction) and quantitative analysis (uptake, engagement with app and wearable, successful use of digital service vs bypass to conventional healthcare contacts).<br>Machine-learning supported exploratory analyses of associations and relative predictive importance of electronic health record, patient-reported outcomes, wearable physiology and NIV parameters: -<br>• Associations between changes in patient-reported outcomes (MRC, CAT, Symptom diary, EQ5-D, S3-NIV and SRF questionnaires) with routine clinical care interventions, COPD exacerbations and other clinical events.<br>• Associations between changes in wearable monitoring parameters (activity, sleep, heart rate variability, energy expenditure, respiratory rate) with routine clinical care interventions, COPD exacerbations and other clinical events.<br>• Associations between changes in NIV monitored parameters (usage, leak, airway pressures, respiratory rate, tidal volume, minute ventilation, inspiratory/expiratory ratio and detected respiratory events) with routine clinical care treatment interventions, COPD exacerbations and other clinical events.<br>• Associations between changes in clinical endpoints and relative importance plots of all remote-management acquired data (including EMG, oscillometry and home pollution monitoring exploratory endpoints in subgroup these measured on) to determine contribution of these to outcome prediction, and therefore value of these for future prospective study.<br>Patient-centred outcomes: -<br>• Health-related quality of life (EQ5-D) at baseline and monthly during study.<br>• Qualitative user research (planned subset of patients, convenience sample) with semi-structured user experience interviews.<br>• Impact of patient activation (where this is measured at baseline and/or follow up during routine clinical care) on enrolment and use of digital service model<br>Healthcare cost analyses: -<br>• Development and installation costs for digitalised service model for remote-management of COPD.<br>• Recurring costs (clinical staffing, digital platform hosting, digital platform scheduled update and maintenance) for digital service model for remote-management of COPD.<br>• Projected direct and indirect cost-savings (admission and unscheduled care reduction, travel, carer-burden impact, |
|  | clinical efficiency) of high-risk COPD with digitally-enabled remote-management, compared with previous service model. |

### Primary endpoint measures

Proportion of patients who engage with digital service model will be determined from screening vs recruitment log, and with patient engagement (PRO completion, wearable monitoring usage, home NIV usage and digital service contacts vs conventional healthcare contacts) monitored using consented platform tracking analytics.

### Secondary endpoint measures

Clinical events, hospital occupied bed days, treatment uptake, NIV usage and therapy, supported self-management success and impact of demographics on these will be captured within the Lenus Health platform with data from weekly SafeHaven export, Trakcare real-time episode data feed and app-platform tracking analytics.

Process evaluation metrics for digital service model will be captured by app-platform tracking analytics, project delivery process documentation and supplementary user experience research conducted as part of the digital service project delivery.

Machine-learning supported analysis of study dataset will be conducted post-hoc.

Patient-centred outcomes will be captured within RECEIVER patient portal. Qualitative user experience and patient activation measurement in planned subgroup (sample of convenience) will be undertaken to inform implementation strategy; this will be presented as a descriptive summary.

Cost analysis will be based on audited accounts for the RECEIVER innovation project delivery, combined with standard NHS tariffs, NSS medical / agenda for change salary scales and indirect costs from standard burden of COPD cost projections.

## Data Management

Study data file will be held in locked cabinet in the Department of Respiratory Medicine, Queen Elizabeth University Hospital, Glasgow, with an electronic copy securely stored on the EDGE Clinical Research Management System. All electronic data and COPD service digital architecture (including machine-learning algorithms) will be held within NHS GG&C e-health systems, with industry standard security and identity assurance processes. The core components of the digital service will be on NHS GG&C Azure cloud tenancy, further ensuring security and identity assurance, and avoid need for data de-identification prior to machine-learning analysis.

Data access will be password protected and accessible only by study investigators, with data management as per NHS GG&C and NHS Scotland data protection policies. Platform analytics will track user interactions and patient data changes, to provide an audit trail for data integrity.

At study completion, the comprehensive study dataset will be submitted for inclusion in NHS GG&C SafeHaven. This will allow (with appropriate SafeHaven SOP and LPAC application approvals) subsequent de-identified review of all study outcomes, re-analysis of the dataset, and contribution of the dataset to future COPD data linkage and other research work within NHS Scotland.

## Statistics and Data Analysis

Secondary outcomes in the RECEIVER observational cohort will be compared with matched retrospective data from de-identified linked datasets of historical control and contemporary control high-risk COPD patients. This will comprise patients identified from coding and admission data as having had a severe exacerbation of COPD between 1st January 2010 - 30th April 2019 (historical cohort) and between 1st July 2019 - 30th June 2020, excluding patients enrolled in the RECEIVER study (contemporary control). The historical control dataset will contribute to machine-learning algorithm for risk predictive models. Secondary outcome analysis will be separately compared between the RECEIVER cohort, historical cohort and contemporary control cohort. This component of the study is also separately registered with dataset access approval from NHS GG&C SafeHaven Local Privacy and Advisory Committee.

Clinical endpoint data will be reported as proportion and 95% CI for that outcome, calculated by Kaplan Meirer method. Demographic data will be presented as mean and standard deviations, or medians and interquartile ranges as appropriate. Correlation, t-test and analysis of variance analyses will be used as appropriate for secondary endpoint analysis comparing results if clinical endpoint and patient-centered outcomes with NIV therapy and supported-self management parameters.

Risk predictive modelling development will be approached as binary classifications with application of machine-learning ensemble methods for development, variation and model performance over time analyses. Precision, recall, accuracy, and C-statistics of the receiver operating characteristic (ROC) curve will be used to evaluate model performances.

## RECEIVER trial qualitative sub-study: Protocol addition and amendment September 2020

Following COVID-19 pandemic, the digital service model established in the RECEIVER trial was adopted for routine clinical care in NHS GG&C. This was to mitigate anticipated COVID-19 impacts on routine COPD care with concern about increased winter admission risks and continued requirement to maintain social distancing with vulnerable/shielding patients. Process for invitation and remote enrolment in the COPD digital service, via support website has been established. Patients are enrolled following clinician referral or via invitations sent by SMS and letter to known patients.

Patient engagement, clinical and service outcomes in this scale-up cohort of patients will be evaluated in parallel with the data from the RECEIVER trial. These analyses will be conducted on de-identified data derived from NHS GG&C SafeHaven. There is separate ethics approval and protocol for these analyses.

It is relevant to gather patient user experience with the remote invitation and enrolment process, with the digital service model and determine whether there are different experiences based on the recruitment source (via clinical trial, via invitation and website registration, via clinician referral).

A randomised sample of convenience of patients who have enrolled in the COPD digital service via this scale-up service model will be established by the clinical team. Patients will be approached by the clinical team via the digital service messaging interface. Patients expressing an interest in undertaking semi-structured interviews about their service experience will be sent RECEIVER sub-study patient information sheet and consent form. These patients would then be contact by one of the study investigators with the consent form discussed and completed by telephone, avoiding COVID-19 context risk of face-face contact for vulnerable patients. Consent form with investigator signature would be mailed to the participant. Semi-structured interviews would then be scheduled and conducted by telephone or video call, using NHS GG&C Attend Anywhere platform.

## Assessment of Safety

The ‘RECEIVER’ digital service model is supporting rather than varying routine clinical care in COPD patients. Adverse and serious adverse events (AEDs and SAEs) will be captured within patient portal (eg PRO question: have you had any hospital admissions?) and within COPD platform (by Trakcare derived clinical events) and provided as a summary report.

Where an SAE requires recording, full details including the nature of the event, start and stop dates, severity, relationship to research product and/or trial procedures, and the outcome of the event will be recorded in the patient’s medical notes and CRFs. These events will be monitored and followed up until satisfactory resolution and stabilisation.

Each SAE will be assessed to determine if related to the research procedures and expectedness where the event is related by the following definitions:

- **Related:** that is, it resulted from administration of study medicines or any of the research procedures
- **Expectedness of SAEs:** is against the research procedure events listed in study protocol as an expected occurrence.

### Expected adverse events

In general, there is little additional risk to participants taking part in the study. However, the study aims to digitise existing reporting pathways and therefore the potential exists for failures to occur within the software used. Expected adverse events related to use of RECEIVER platform are as follows:

- Data connectivity issues: There may be issues with transfer of data from the patient client to REVCEIVER platform. Where this data relates to symptoms diaries or Fitbit wearable data there would be no impact on clinical care should the loss of data be temporary. However, in the event the data was permanently lost this may impact on patient care.
- Messaging: There is a potential for messages sent from the patient to study staff (and vice versa) via the RECEIVER platform to be missed. There is also the potential for these messages to be confusing to the patient.
- Identity: There is the potential for data for one patient to be allocated incorrectly to another patient due to errors within the RECEIVER platform software.

Adverse effects related to the use of the REVEIVER platform (AEDs) are not considered reportable to the Sponsor.

Exacerbations of COPD resulting in hospitalisation and potentially death are expected within the study population. In addition, patients with COPD may have other comorbid conditions. Events related to these conditions that meet the criteria of an SAE would also be considered expected. SAEs related to the participants underlying medical condition(s) that are not causally related to the RECIEVER platform are not reportable to the Sponsor.

Safety Reporting to Sponsor

The following events are reportable to the Sponsor:

- Any SAE that is causally related to the use of RECEIVER platform (Serious Adverse Device Effect – SADE) regardless or expectedness.
- Any SAE that is related to the patient’s participation within the trial that is both related and unexpected but is not related to the use of the RECEIVER platform (Related and Unexpected Serious Adverse Effect – RUSAEs)

Study related unexpected SAE (SRU-SAEs) must be reported to the Pharmacovigilance (PV) office immediately (within 24 hours). The SAE form should be completed and signed by appropriately delegated staff. If necessary, a verbal report can be given by contacting the PV office. This must be followed up as soon as possible with a signed written (or electronic) report. If all of the required information is not available at the time of initial reporting, the CI (or designee) must ensure that any missing information is forwarded to the PV office as soon as this becomes available. The report should indicate that this information is follow-up information for a previously reported event.

The PV office will report all RUSAEs and unexpected SADEs (USADEs) to the ethics committee within 15 days of the PV office becoming aware of the event, via the ‘report of serious adverse event form’ for non-CTIMPs published on the Health Research Authority web site.

RUSAEs and USADEs will also be considered individually by project steering group. Where appropriate modifications (e.g. additional patient alert notifications, additional clinical decision support notifications) to the RECEIVER platform will be made, in discussion with ethics committee and supported by protocol amendment.

Annual progress / safety report will be provided by the Chief Investigator to the REC and R&D department.

### Protocol Amendments

Any change in the study protocol will require an amendment. Any proposed protocol amendments will be initiated by the PI following discussion with the project steering group. Any required amendments forms will be submitted to the regulatory authority, ethics committee and Sponsor. The PI and the project steering group will liaise with study Sponsor to determine whether an amendment is non-substantial or substantial. All amended versions of the protocol will be signed by PI and Sponsor representative. Before the amended protocol can be implemented favourable opinion/approval must be sought from the original reviewing REC and Research and Development (R&D) office(s).

## Ethics and Dissemination

This study will be carried out in accordance with the World Medical Association Declaration of Helsinki (1964) and its revisions (Tokyo [1975], Venice [1983], Hong Kong [1989], South Africa [1996] and Edinburgh [2000]).

Ethical approval for this clinical trial has been obtained from the West of Scotland Research Ethics Service (WoSRES). Patients will only be allowed to enter the study once they have provided written informed consent. This CI will be responsible for updating the Ethics committee of any new information related to the study.

Key results will be presented at local, national and international meetings, including those with patient representation. All data obtained will be submitted for publication to peer reviewed journals. Principle and co-investigators will have access to all data analyses conducted by project commercial partners (StormID), with investigators having full academic independence for publication of results.

The study has been registered with clinicaltrials.gov (NCT04240353).

## Data Availability

Data obtained from the RECEIVER trial is available for NHS GG&C SafeHaven.

https://support.nhscopd.scot

## Funding and Indemnity

This study is sponsored by NHS Greater Glasgow & Clyde. The sponsor will be liable for negligent harm caused by the design of the trial. NHS indemnity is provided under Clinical Negligence and Other Risks Indemnity Scheme (CNORIS).

This work was supported by Innovate UK, grant number/project ID 104552

## Authors contribution

I - Conception and design: PM, CC, DL, GM

II - Administrative support: All

III - Provision of study material or patients: n/a

IV - Collection and assembly of data: n/a

V - Data analysis and interpretation: n/a

VI - Manuscript writing: AT, GM, PM, DL, CC

VII - Final approval of manuscript: All

## Appendix 1: RECEIVER COPD Patient-web portal questionnaires

### COPD Patient App v5.0

Patient Reported Outcome Flows

Daily

#### Symptom diary

1. How are you feeling today? (1) Better than usual (2) Normal/usual (3) Worse than usual (4) Much worse than usual
2. How is your breathing today? (1) Better than usual (2) Normal/usual (3) Worse than usual (4) Much worse than usual
3. Do you have a cold or flu today?
  - Yes
  - No

#### CAT (score/40)

4. (0) I never cough (1) (2) (3) (4) (5) I cough all the time
5. (0) I have no phlegm (mucus) in my chest at all (1) (2) (3) (4) (5) My chest is completely full of phlegm (mucus)

#### Symptom diary additional questions

How difficult is it to bring up phlegm when you cough?

(1) Not difficult

(2) A little difficult

(3) Quite difficult

(4) Very difficult

What consistency is your phlegm?

(1) Watery

(2) Sticky liquid

(3) Semi-solid

(4) Solid

What colour is your phlegm?

(1) White

(2) Yellow

(3) Green

(4) Dark green

6. (0) My chest does not feel tight at all (1) (2) (3) (4) (5) My chest feels very tight
7. (0) When I walk up a hill or one flight of stairs I am not breathless (1) (2) (3) (4) (5) When I walk up a hill or one flight of stairs I am very breathless
8. (0) I am not limited doing any activities at home (1) (2) (3) (4) (5) I am very limited doing activities at home
9. (0) I am confident leaving my home despite my lung condition (1) (2) (3) (4) (5) I am not at all confident leaving my home because of my lung condition
10. (0) I sleep soundly (1) (2) (3) (4) (5) I don’t sleep soundly because of my lung condition
11. (0) I have lots of energy (1) (2) (3) (4) (5) I have no energy at all

Weekly

#### Symptom diary

1. How are you feeling today? (1) Better than usual (2) Normal/usual (3) Worse than usual (4) Much worse than usual
2. How is your breathing today? (1) Better than usual (2) Normal/usual (3) Worse than usual (4) Much worse than usual
3. Do you have a cold or flu today?
  - Yes
  - No
4. Have you increased your usual breathing treatment this week? (e.g. inhalers, nebulisers, tablets)
  - Yes
  - No
5. Have you taken antibiotics this week?
  - Yes
  - No
6. Have you visited your GP this week?
  - Yes
  - No
7. Have you visited your hospital doctor this week?
  - Yes
  - No

#### CAT (score /40) (For each of the following questions, please select the number that best describes you currently.)

8. (0) I never cough (1) (2) (3) (4) (5) I cough all the time
9. (0) I have no phlegm (mucus) in my chest at all (1) (2) (3) (4) (5) My chest is completely full of phlegm (mucus)

#### Symptom diary additional questions

How difficult is it to bring up phlegm when you cough?

- Not difficult
- A little difficult
- Quite difficult
- Very difficult

What consistency is your phlegm?

- Watery
- Sticky liquid
- Semi-solid
- Solid

What colour is your phlegm?

- White
- Yellow
- Green
- Dark green

10. (0) My chest does not feel tight at all (1) (2) (3) (4) (5) My chest feels very tight
11. (0) When I walk up a hill or one flight of stairs I am not breathless (1) (2) (3) (4) (5) When I walk up a hill or one flight of stairs I am very breathless
12. (0) I am not limited doing any activities at home (1) (2) (3) (4) (5) I am very limited doing activities at home
13. (0) I am confident leaving my home despite my lung condition (1) (2) (3) (4) (5) I am not at all confident leaving my home because of my lung condition
14. (0) I sleep soundly (1) (2) (3) (4) (5) I don’t sleep soundly because of my lung condition
15. (0) I have lots of energy (1) (2) (3) (4) (5) I have no energy at all

#### MRC (score /4)

Please tick in the box that applies to you (one box only):

1. I only get breathless with strenuous exercise
1. I get short of breath when hurrying on the level or walking up and slight hill
2. I walk slower than people of the same age on the level because of breathlessness or have to stop for breath when walking at my own pace on the level
3. I stop for breath after walking about 100 yards or after a few minutes on the level
4. I am too breathless to leave the house or I am breathless when dressing

Every 4^th^ week

#### Symptom diary

16. How are you feeling today? (1) Better than usual (2) Normal/usual (3) Worse than usual (4) Much worse than usual
17. How is your breathing today? (1) Better than usual (2) Normal/usual (3) Worse than usual (4) Much worse than usual
18. Do you have a cold or flu today?
  - Yes
  - No
19. Have you increased your usual breathing treatment this week? (e.g. inhalers, nebulisers, tablets)
  - Yes
  - No
20. “Have you taken a rescue pack or an acute course of antibiotics or steroids prescribed by a doctor for your COPD this week? *This does not include long-term antibiotics*.*”?*
  - Yes
  - No
21. Have you visited your GP this week?
  - Yes
  - No
22. Have you visited your hospital doctor this week?
  - Yes
  - No

#### CAT (score /40) (For each of the following questions, please select the number that best describes you currently.)

23. (0) I never cough (1) (2) (3) (4) (5) I cough all the time
24. (0) I have no phlegm (mucus) in my chest at all (1) (2) (3) (4) (5) My chest is completely full of phlegm (mucus)

#### Symptom diary additional questions

How difficult is it to bring up phlegm when you cough?

- Not difficult
- A little difficult
- Quite difficult
- Very difficult

What consistency is your phlegm?

- Watery
- Sticky liquid
- Semi-solid
- Solid

What colour is your phlegm?

- White
- Yellow
- Green
- Dark green

25. (0) My chest does not feel tight at all (1) (2) (3) (4) (5) My chest feels very tight
26. (0) When I walk up a hill or one flight of stairs I am not breathless (1) (2) (3) (4) (5) When I walk up a hill or one flight of stairs I am very breathless
27. (0) I am not limited doing any activities at home (1) (2) (3) (4) (5) I am very limited doing activities at home
28. (0) I am confident leaving my home despite my lung condition (1) (2) (3) (4) (5) I am not at all confident leaving my home because of my lung condition
29. (0) I sleep soundly (1) (2) (3) (4) (5) I don’t sleep soundly because of my lung condition
30. (0) I have lots of energy (1) (2) (3) (4) (5) I have no energy at all

#### MRC (score /4)

Please tick in the box that applies to you (one box only):

1. I only get breathless with strenuous exercise
1. I get short of breath when hurrying on the level or walking up and slight hill
2. I walk slower than people of the same age on the level because of breathlessness or have to stop for breath when walking at my own pace on the level
3. I stop for breath after walking about 100 yards or after a few minutes on the level
4. I am too breathless to leave the house or I am breathless when dressing

Every month we are asking some additional research questions to help us understand the impact of COPD on your quality of life. These questions will take a further couple of minutes, but can be skipped if you prefer *(with opt in/out click option)*

#### Quality of Life (EQ5D)

Mobility

- I have no problems in walking about
- I have slight problems in walking about
- I have moderate problems in walking about
- I have severe problems in walking about
- I am unable to walk about

Self-care

- I have no problems washing or dressing myself
- I have slight problems washing or dressing myself
- I have moderate problems washing or dressing myself
- I have severe problems washing or dressing myself
- I am unable to wash or dress myself

Usual activities (e.g. work, study, housework, family or leisure activities)

- I have no problems doing my usual activities
- I have slight problems doing my usual activities
- I have moderate problems doing my usual activities
- I have severe problems doing my usual activities
- I am unable to do my usual activities.

Pain/discomfort

- I have no pain or discomfort
- I have slight pain or discomfort
- I have moderate pain or discomfort
- I have severe pain or discomfort
- I have extreme pain or discomfort

Anxiety/depression

- I am not anxious or depressed
- I am slightly anxious or depressed
- I am moderately anxious or depressed
- I am severely anxious or depressed
- I am extremely anxious or depressed

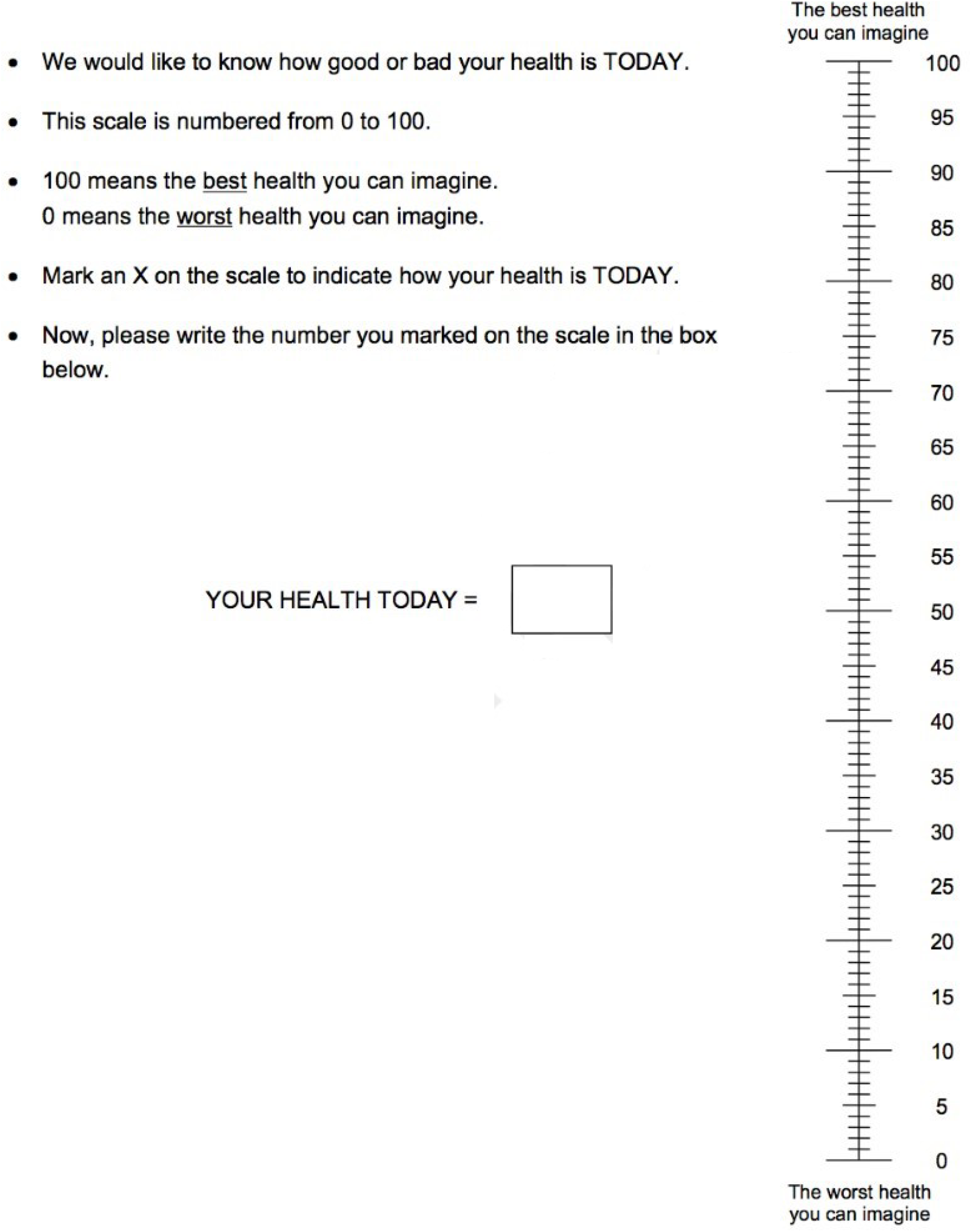

## Appendix 2: RECEIVER Trial – Digitising Routine COPD care

The components of the RECEIVER trial are a digitisation of routine clinical care. All of these components, including integrated care and supported self-management are as recommended in international consensus guidelines for COPD management: Global Obstructive Lung Disease (GOLD) guideline 2019.

Table below lists the trial schedule / study components, summarising current routine care and how this is digitised.

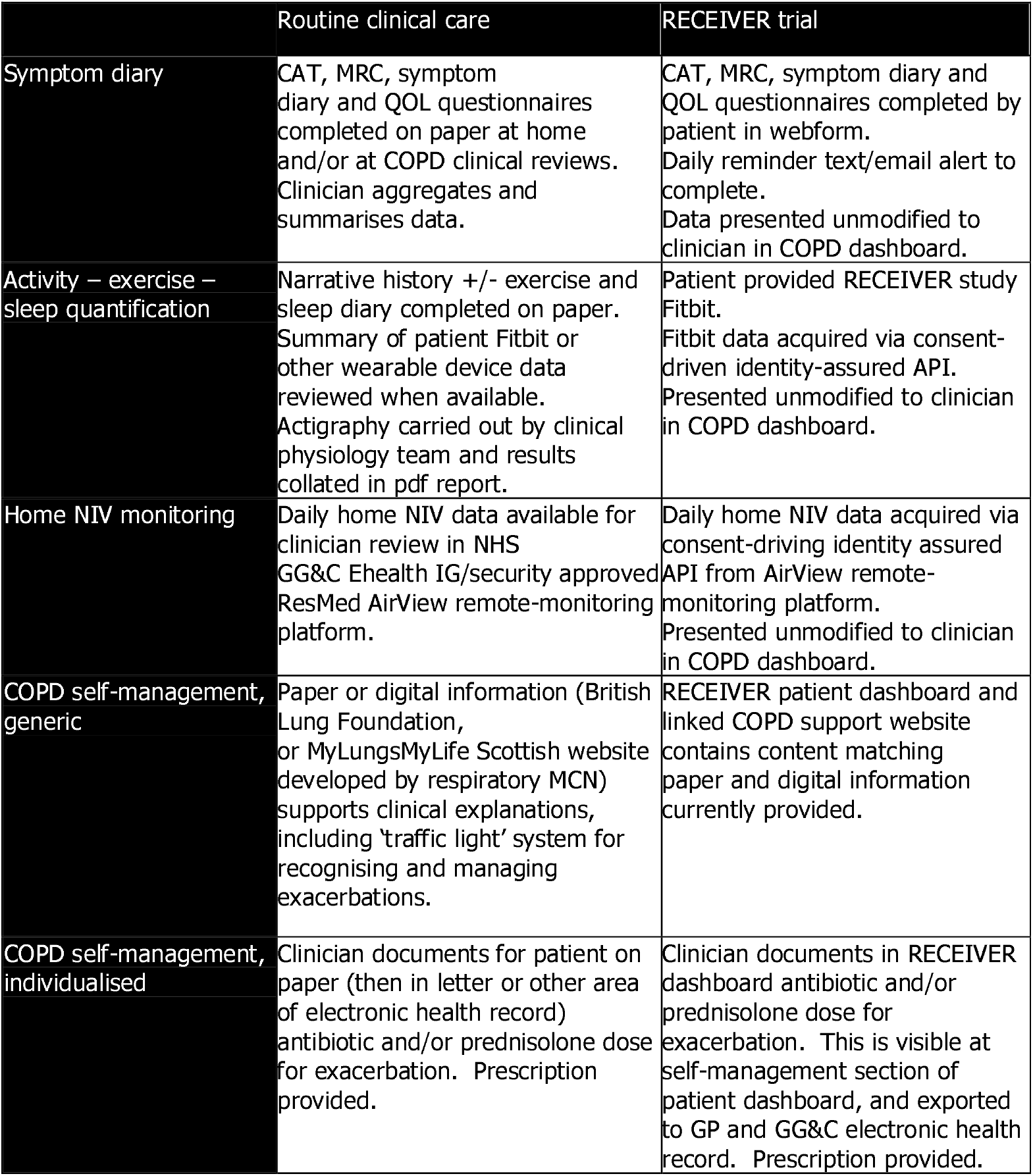
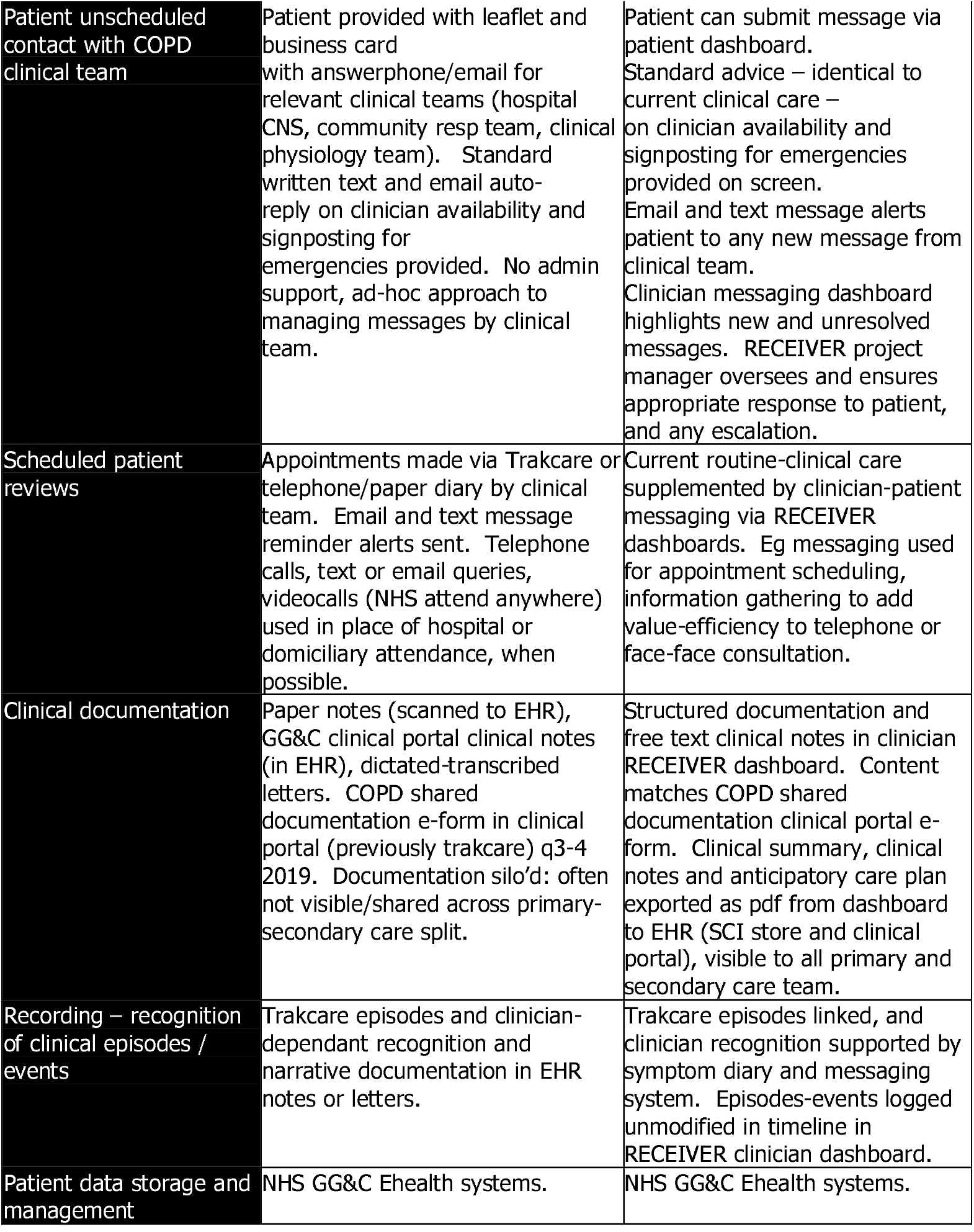

## Appendix 3: RECEIVER Trial – Data Storage in NHS GG&C Azure tenancy

The working components of the COPD digital service are maintained within the NHS GG&C Lenus account. The databases containing the patient data from the Receiver trial, and the historical and contemporary control cohort data from NHS GG&C SafeHaven is maintained in a separate account, with restricted access. Planned analyses of these datasets is subject to LPAC approvals, and SafeHaven SOPs to ensure only de-identified data is shared.

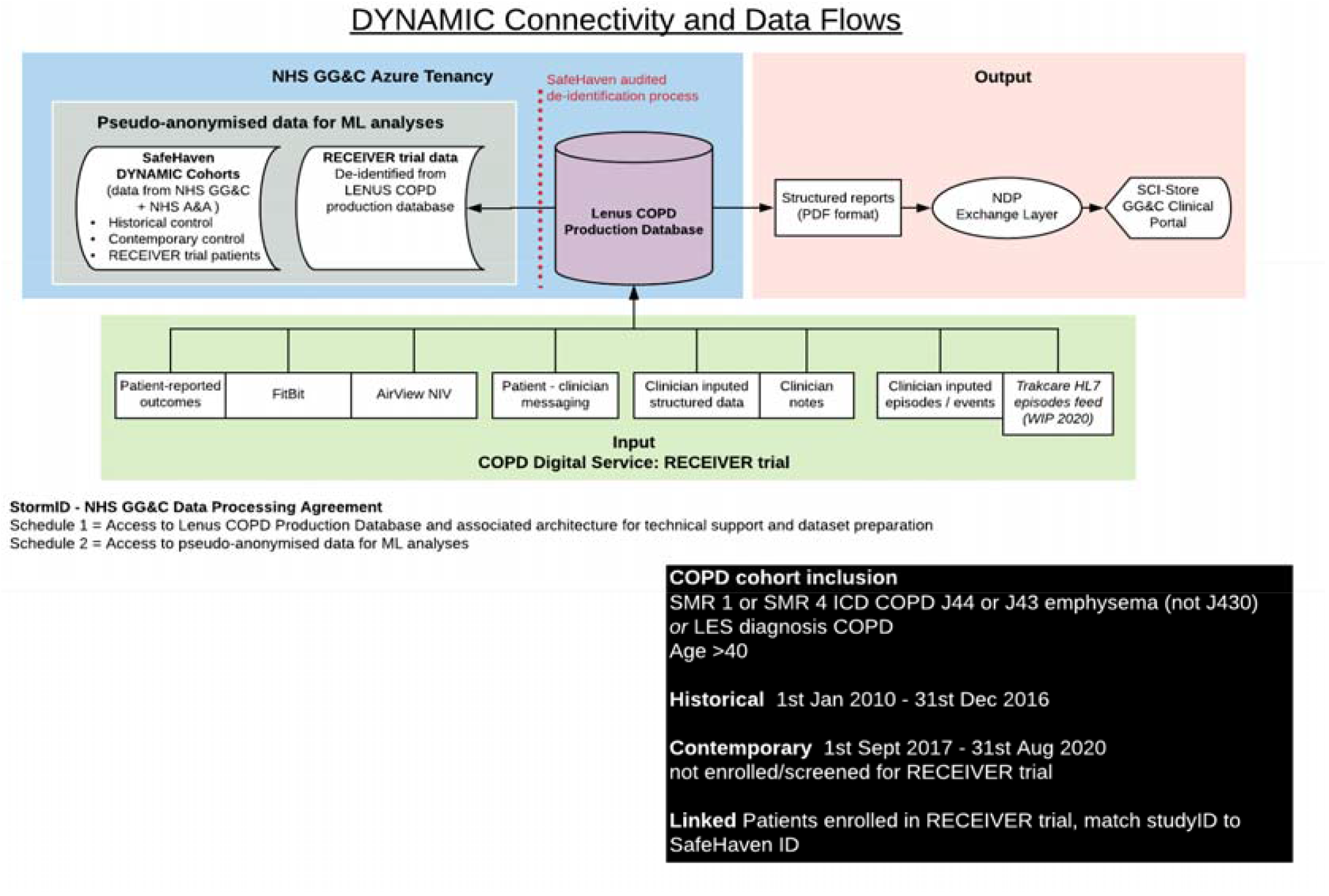

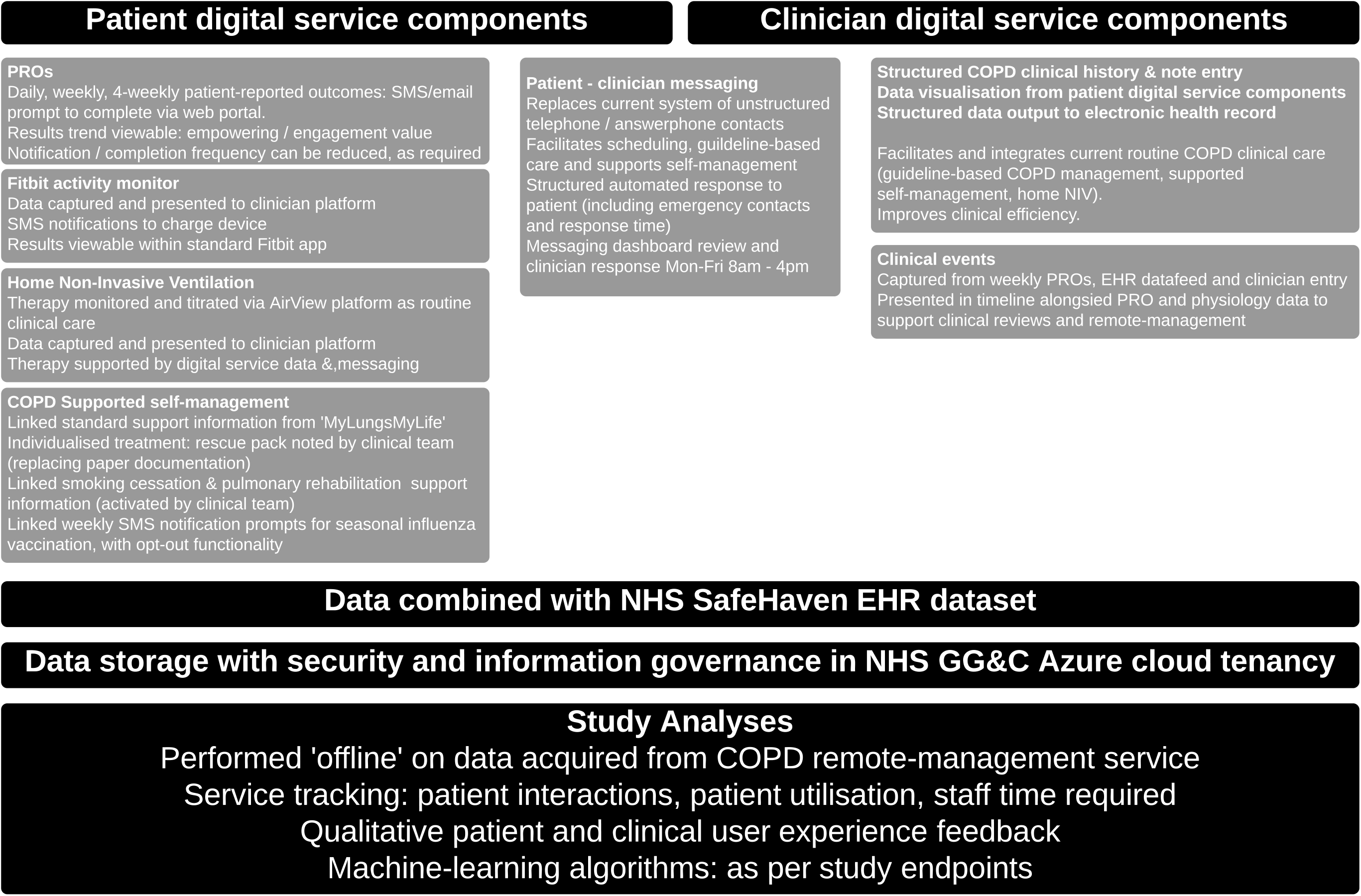

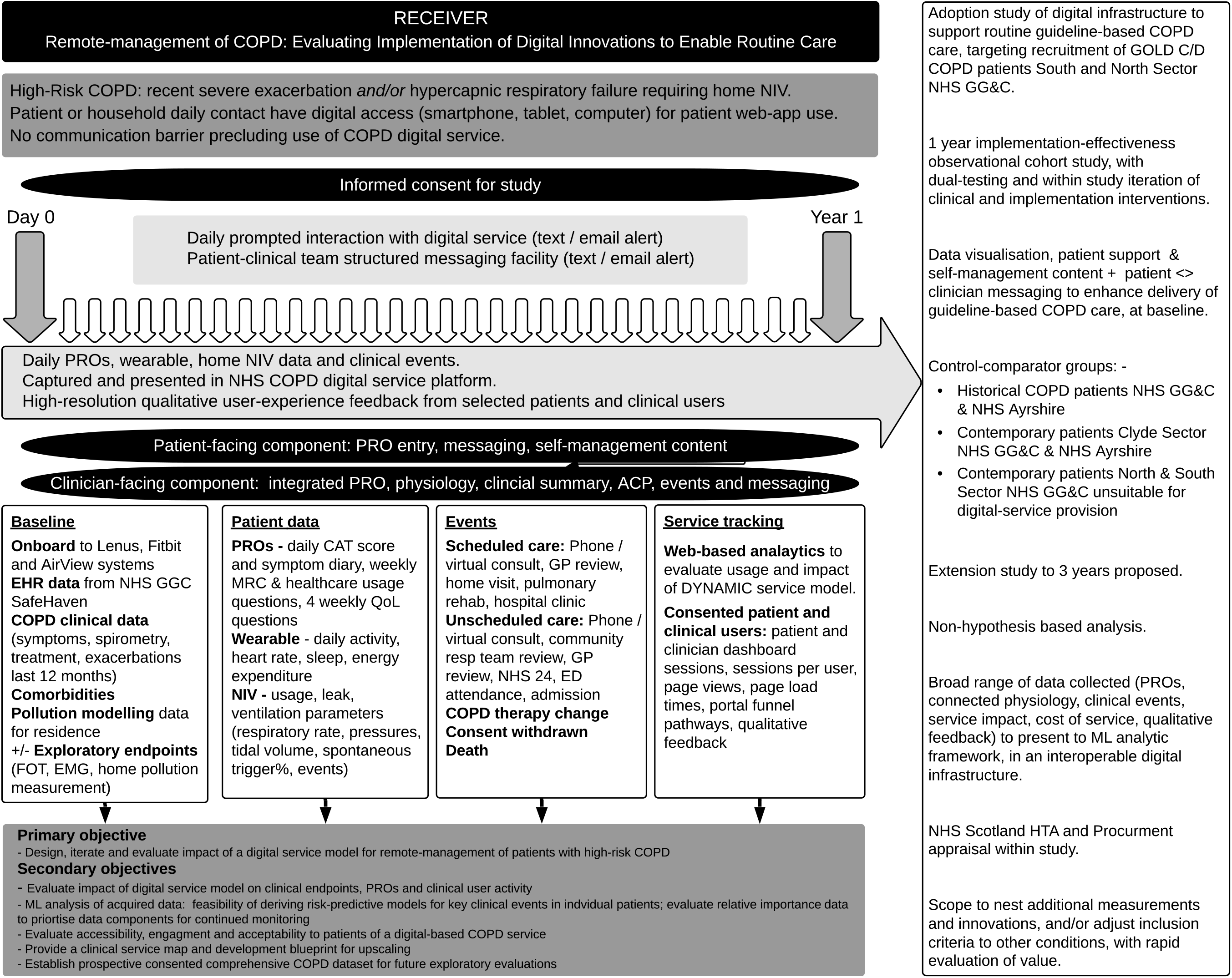

