## Supplementary material 1 for "Remote-management of COPD: Evaluating Implementation of Digital Innovation to Enable Routine Care (RECEIVER) – Protocol for a feasibility and service adoption observational cohort study"

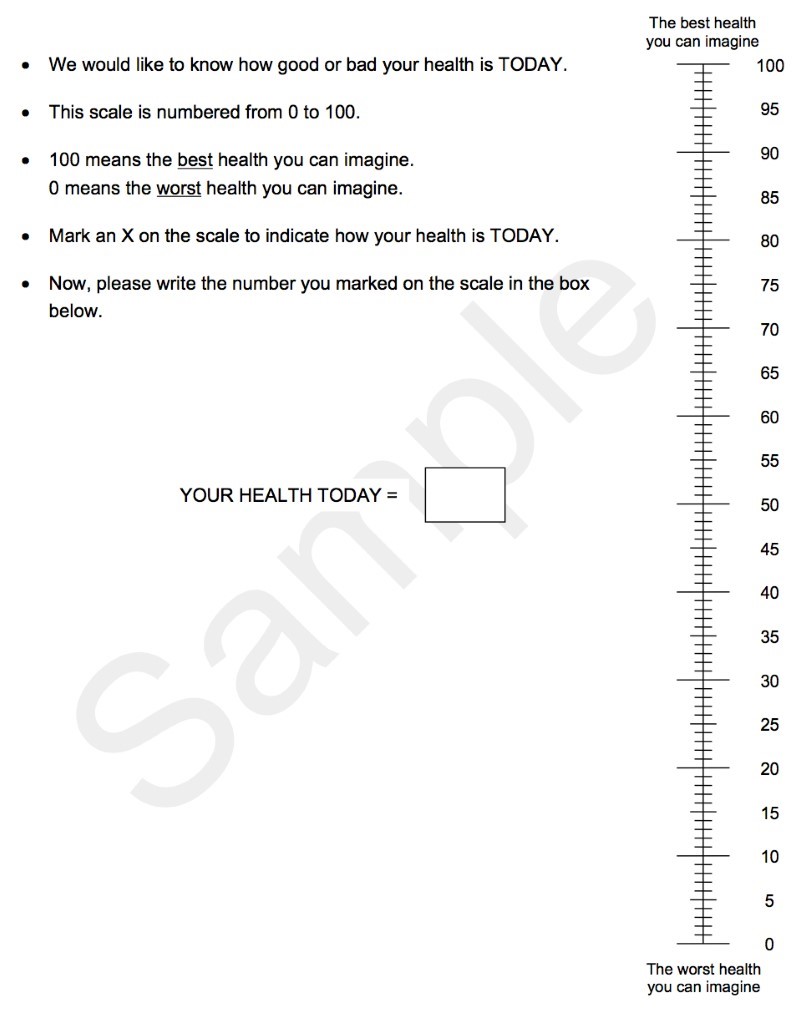
