## Supplementary material 2 for "Remote-management of COPD: Evaluating Implementation of Digital Innovation to Enable Routine Care (RECEIVER) – Protocol for a feasibility and service adoption observational cohort study"

**Supplementary material 2: RECEIVER Trial – Digitising Routine COPD care**

The components of the RECEIVER trial are a digitisation of routine clinical care.  All of these components, including integrated care and supported self-management are as recommended in international consensus guidelines for COPD management: Global Obstructive Lung Disease (GOLD) guideline 2019.

Table below lists the trial schedule / study components, summarising current routine care and how this is digitised.

|  | Routine clinical care | RECEIVER trial |
| --- | --- | --- |
| Symptom diary | CAT, MRC, symptom diary and QOL questionnaires completed on paper at home and/or at COPD clinical reviews.  Clinician aggregates and summarises data. | CAT, MRC, symptom diary and QOL questionnaires completed by patient in webform.    Daily reminder text/email alert to complete.     Data presented unmodified to clinician in COPD dashboard. |
| Activity – exercise – sleep quantification | Narrative history +/- exercise and sleep diary completed on paper.  Summary of patient Fitbit or other wearable device data reviewed when available.  Actigraphy carried out by clinical physiology team and results collated in pdf report. | Patient provided RECEIVER study Fitbit.    Fitbit data acquired via consent-driven identity-assured API.    Presented unmodified to clinician in COPD dashboard. |
| Home NIV monitoring | Daily home NIV data available for clinician review in NHS GG&C Ehealth IG/security approved ResMed AirView remote-monitoring platform. | Daily home NIV data acquired via consent-driving identity assured API from AirView remote-monitoring platform.  Presented unmodified to clinician in COPD dashboard. |
| COPD self-management, generic | Paper or digital information (British Lung Foundation, or MyLungsMyLife Scottish website developed by respiratory MCN) supports clinical explanations, including ‘traffic light’ system for recognising and managing exacerbations. | RECEIVER patient dashboard and linked COPD support website contains content matching paper and digital information currently provided. |
| COPD self-management, individualised | Clinician documents for patient on paper (then in letter or other area of electronic health record) antibiotic and/or prednisolone dose for exacerbation.  Prescription provided. | Clinician documents in RECEIVER dashboard antibiotic and/or prednisolone dose for exacerbation.  This is visible at self-management section of patient dashboard, and exported to GP and GG&C electronic health record.  Prescription provided. |
| Patient unscheduled contact with COPD clinical team | Patient provided with leaflet and business card with answerphone/email for relevant clinical teams (hospital CNS, community resp team, clinical physiology team).   Standard written text and email auto-reply on clinician availability and signposting for emergencies provided.  No admin support, ad-hoc approach to managing messages by clinical team. | Patient can submit message via patient dashboard.  Standard advice – identical to current clinical care – on clinician availability and signposting for emergencies provided on screen.   Email and text message alerts patient to any new message from clinical team.  Clinician messaging dashboard highlights new and unresolved messages.  RECEIVER project manager oversees and ensures appropriate response to patient, and any escalation. |
| Scheduled patient reviews | Appointments made via Trakcare or telephone/paper diary by clinical team.  Email and text message reminder alerts sent.  Telephone calls, text or email queries, videocalls (NHS attend anywhere) used in place of hospital or domiciliary attendance, when possible. | Current routine-clinical care supplemented by clinician-patient messaging via RECEIVER dashboards.  Eg messaging used for appointment scheduling, information gathering to add value-efficiency to telephone or face-face consultation. |
| Clinical documentation | Paper notes (scanned to EHR), GG&C clinical portal clinical notes (in EHR), dictated-transcribed letters.  COPD shared documentation e-form in clinical portal (previously trakcare) q3-4 2019.  Documentation silo’d: often not visible/shared across primary-secondary care split. | Structured documentation and free text clinical notes in clinician RECEIVER dashboard.  Content matches COPD shared documentation clinical portal e-form.  Clinical summary, clinical notes and anticipatory care plan exported as pdf from dashboard to EHR (SCI store and clinical portal), visible to all primary and secondary care team. |
| Recording – recognition of clinical episodes / events | Trakcare episodes and clinician-dependant recognition and narrative documentation in EHR notes or letters. | Trakcare episodes linked, and clinician recognition supported by symptom diary and messaging system.  Episodes-events logged unmodified in timeline in RECEIVER clinician dashboard. |
| Patient data storage and management | NHS GG&C Ehealth systems. | NHS GG&C Ehealth systems. |
