## Supplementary material 3 for "Remote-management of COPD: Evaluating Implementation of Digital Innovation to Enable Routine Care (RECEIVER) – Protocol for a feasibility and service adoption observational cohort study"

**Supplementary material 3: RECEIVER Trial – Data Storage in NHS GG&C Azure tenancy**

The working components of the COPD digital service are maintained within the NHS GG&C Lenus account.  The databases containing the patient data from the Receiver trial, and the historical and contemporary control cohort data from NHS GG&C SafeHaven is maintained in a separate account, with restricted access.  Planned analyses of these datasets is subject to LPAC approvals, and SafeHaven SOPs to ensure only de-identified data is shared.

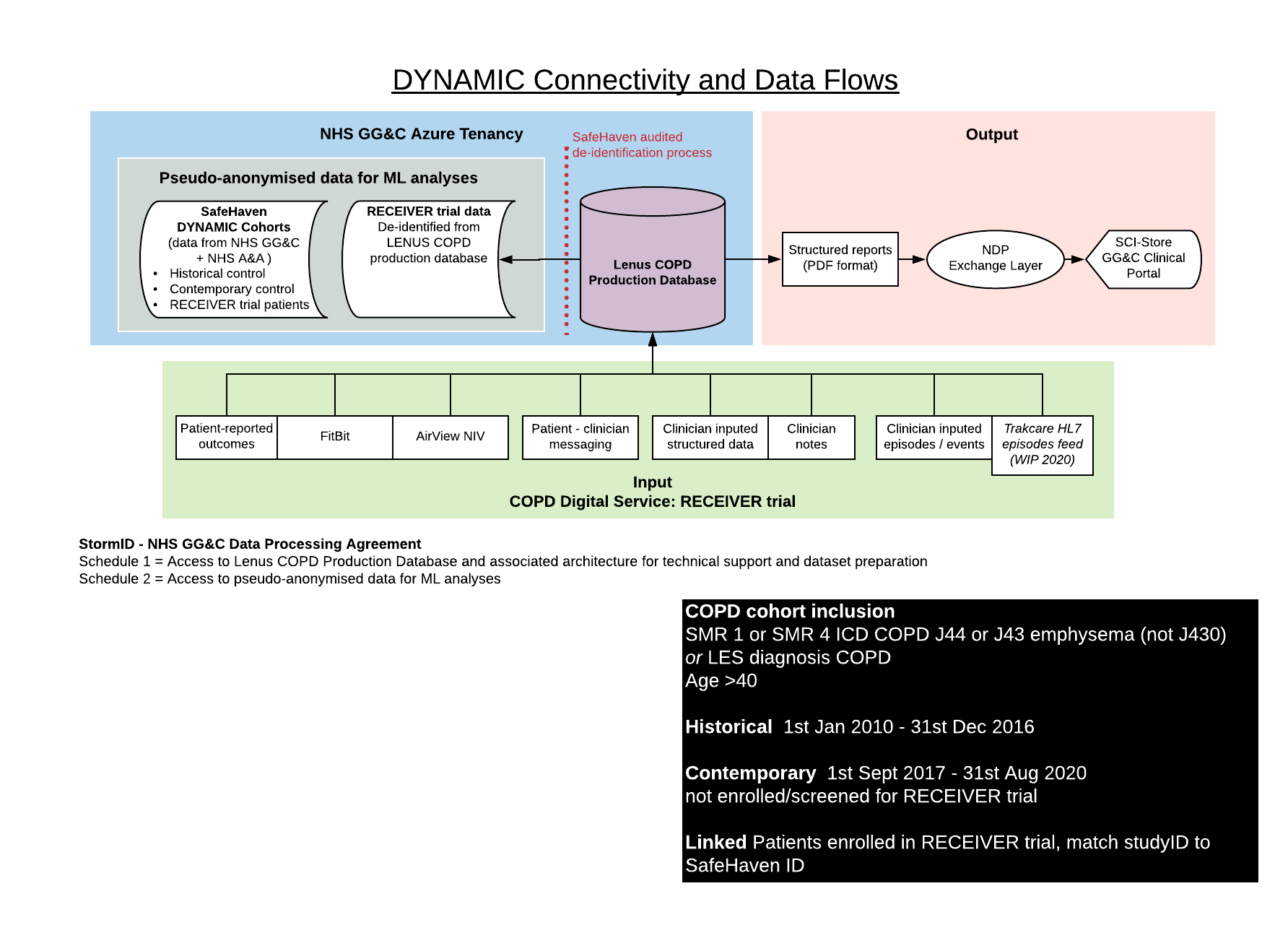
